# MinderCare: protocol for a mixed-methods evaluation of a digitally enabled dementia care service

**DOI:** 10.64898/2026.06.19.26356077

**Authors:** Julian Jeyasingh-Jacob, Margherita Tecilla, Suzie Cro, Helen Lai, Gaia Frigerio, Nora Joby, Catalina Chavarro Novoa, Success Fabusoro, Maria Rasulo, Joanna James, Janibo Amade Cassimo, Farah Harris, Mara Golemme, Tori Simpson, Matthew Harrison, Ebenezar Ndachi Effiang, CR&T Group, Sarah Daniels, Eyal Soreq, David J Sharp

**Author notes:** These authors made equal first author contributions (Julian Jeyasingh-Jacob, Margherita Tecilla). These authors made equal senior author contributions (Eyal Soreq, David J Sharp).

## Abstract

**Introduction and aims:** Dementia is a growing public health challenge affecting millions of people worldwide. It is a progressive condition that increases the risk of infections, falls, hospital admissions, dependence in activities of daily living, safety issues such as wandering, care home transfers, and death. New ways of supporting people living with dementia (PLWD) at home are urgently needed. We describe the MinderCare study which evaluates a digitally enabled care model that integrates low-burden sensor-based remote monitoring within a nurse-led clinical service.

**Methods and analysis:** In this mixed-methods study, we will recruit 100 people with confirmed or suspected dementia living at home and deploy the Minder remote monitoring system for at least 12 months. A detailed characterisation of the cohort will be obtained, including cognition, frailty, participant and carer wellbeing, functioning, and quality of life. The feasibility, acceptability, sustainability, and resource requirements of the service will also be assessed. Low-cost sensors provide information about behaviour, environment and physiology from the home. Machine-learning algorithms have been used to develop digital biomarkers of infection, sleep, night-time behaviours, daily activities and routines, and the effects of clinical events and treatment. These will be assessed through clinical reports of sensor-derived data that include anomaly alerts provided to the clinical teams. Algorithms will be assessed for their clinical utility and acceptability. The comparative-effectiveness component will be designed as a target trial emulation using linked electronic health-record data to construct a time-indexed external usual-care control cohort. The primary comparative outcome will be Days Alive and Out of Hospital (DAOH) over 12 months from the activation-index date, with healthcare utilisation, costs, institutionalisation and mortality assessed as secondary outcomes. DAOH and estimated MinderCare effects will also be examined across prespecified strata of baseline inpatient utilisation.

**Ethics and dissemination:** Ethical approval has been granted by the North East – Newcastle and North Tyneside 2 Research Ethics Committee, and the study has received confirmation of capacity and capability by the Imperial College Healthcare NHS Trust. Study findings will be disseminated to patients, health and social care professionals, and policymakers through peer-reviewed publications and conference presentations.

Study registration number: ISRCTN14997677 and NIHR portfolio CPMSID 63023.

**Strengths and limitations of this study:**

- This study evaluates a digitally enabled remote monitoring model for people living with dementia that integrates passive in-home sensors and algorithm-derived digital biomarkers to detect clinically relevant changes such as infection risk, behavioural disturbance and physiological deterioration.
- The study is embedded within the North West London Integrated Care System, allowing evaluation of a remote monitoring service implemented within routine NHS and social care pathways in a large and socio-demographically diverse population.
- The mixed-methods design integrates continuous sensor data, standardised clinical assessments, linked electronic health records and qualitative interviews to examine algorithm performance alongside service feasibility, acceptability and sustainability.
- The comparative-effectiveness component is structured as a target trial emulation using linked electronic health records, with explicit specification of eligibility, treatment strategies, activation-index date, follow-up, outcomes, causal estimand and analysis plan.
- As a non-randomised target trial emulation using an externally matched comparator, the study may remain susceptible to residual confounding, particularly from unmeasured factors such as carer engagement, home suitability, willingness to accept monitoring and referral-route effects; attrition due to death, care home transition, and variability in home environments or device connectivity may also affect data completeness and interpretation.

## (A) INTRODUCTION

Dementia is one of the most significant global public health and social care challenges of the 21st century. It is a common, progressive neurodegenerative condition affecting an estimated 55 million people worldwide, with annual societal costs exceeding US$1.3 trillion [1]. In the UK, almost one million people are living with dementia, generating an annual economic burden of approximately £42 billion [2]. As prevalence rises, a key challenge is how to identify deterioration earlier and deliver timely community-based support that helps people living with dementia (PLWD) remain safely at home and avoid preventable crises.

Here we describe the MinderCare study, which evaluates digitally enabled dementia care embedded within a nurse-led NHS dementia service. The study builds on prior work from the UK Dementia Research Institute Care Research and Technology Centre (UK DRI CR&T), which has developed and evaluated the Minder platform over the past seven years. Minder is a digitally enabled system for continuous, unobtrusive in-home monitoring in PLWD. Previous Minder studies have demonstrated proof-of-principle for detecting early signals associated with infection, agitation, social isolation, sleep disturbance, and treatment response [3–9]

The approach aligns with current NHS ambitions to shift care from hospital to home and expand the use of remote monitoring and digital decision support in long-term condition management [10]. In MinderCare, this policy context is relevant because it supports implementation, but the key question remains whether such a model is workable, acceptable, and beneficial in routine dementia care.

The impact of dementia extends beyond its direct symptoms. PLWD experience progressive loss of independence and quality of life [11] and are highly vulnerable to acute health crises. In England, a substantial proportion of hospital care is delivered to PLWD, and many admissions are potentially avoidable and associated with infections, falls, or neuropsychiatric complications [12–14]. Hospitalisation is often distressing and disorientating for PLWD and is associated with cognitive and functional decline and increased mortality risk [15–18]. At the same time, dementia places sustained demands on informal carers and families. Caregiver strain is a major contributor to the breakdown of community-based support and subsequent long-term institutionalisation [19]. Together, these pressures highlight the importance of identifying potentially reversible deterioration early, including changes consistent with infection, sleep disturbance, behavioural change, reduced activity, or disruption of daily routines, before they escalate into crisis.

Most PLWD prefer to remain safely and independently in their own homes for as long as possible [20]. However, current health and social care systems often struggle to provide the coordinated, proactive support required to sustain community living. In practice, care is frequently episodic, visit-based, and dependent on deterioration already being visible to carers or clinicians. As a result, subtle day-to-day changes may be missed until minor problems escalate into crises, leading to emergency healthcare use and hospital admission [21,22].

Reducing avoidable deterioration and delaying long-term institutional care are major clinical, service, and economic priorities [23]. Structured home-based support may reduce institutionalisation by as much as 22% [24]. However, it remains unclear whether home-based digital monitoring embedded within routine NHS care pathways can identify deterioration early enough to support timely intervention, reduce crisis-driven care, and help sustain community living.

MinderCare assesses the translational step from platform development to service delivery. It integrates the Minder platform within NHS services to support proactive, person-centred care through continuous passive monitoring, clinical interpretation, and coordinated response. The model of care is intended to reduce reliance on reactive and fragmented services, provide more timely support to carers, and help PLWD remain safely at home. At the same time, these anticipated benefits require evaluation in routine care rather than assumption from proof-of-principle platform studies alone.

Digital support for dementia care is most useful when it can capture changes in health and behaviour continuously and unobtrusively in real-world home environments. Passive sensor systems are particularly well suited to this setting because they do not rely on sustained user interaction, can generate continuous longitudinal data with low participant burden, and may be more scalable and acceptable for people living with cognitive, language, or physical limitations [25,26]. MinderCare uses passive sensor-derived insights within NHS care pathways to support earlier detection of clinically relevant change and more timely responses.

Evidence from coordinated community dementia care programmes further supports the value of proactive, multidisciplinary support. Such models have been associated with reduced long-term care admissions and shorter hospital stays, although effects on overall hospital use have been mixed [27–30]. However, most existing programmes have relied on scheduled review and care coordination rather than continuous home monitoring and real-time sensor-informed alerting. MinderCare is designed to extend these approaches by combining coordinated care with continuous passive monitoring to identify deterioration earlier and support more timely action.

This study is conducted within the Northwest London Integrated Care System, which serves approximately 2.4 million people across eight London boroughs and includes substantial ethnic and socioeconomic diversity, marked health inequalities, and a high burden of long-term conditions, including dementia [31]. This setting is methodologically important because it provides a real-world integrated care environment in which MinderCare can be evaluated at scale, while linked regional health and care data also support the construction of an electronically matched external comparator [32].

Scaling community dementia support is challenged by workforce constraints and the resource-intensive nature of traditional visit-based monitoring[33,34]. Integrating sensor data into shared-care platforms may support coordinated multidisciplinary workflows and allow more scalable monitoring models, including centralised oversight of larger caseloads [35]. However, digital inequities remain a major concern, particularly for older adults facing barriers related to connectivity, device access, and digital literacy [36]. These considerations make it essential to evaluate not only clinical and comparative outcomes but also feasibility, acceptability, sustainability, and equity of implementation.

This protocol describes a mixed-methods evaluation of MinderCare, a digitally enabled dementia care model that combines passive in-home monitoring with clinical review and coordinated response. The study comprises linked components including a prospective evaluation of sensor-derived digital biomarkers against clinical ground-truth data; an assessment of service delivery, user experience, sustainability, and implementation within routine NHS and social care pathways; a comparative effectiveness evaluation; and a health economic evaluation using an electronically matched external control cohort derived from the Discover-NOW linked health and care dataset for Northwest London. For each MinderCare participant, candidate controls will be drawn from the contemporaneously eligible population and matched on baseline demographic and clinical characteristics, including age, sex, ethnicity, deprivation, dementia subtype, frailty, and prior healthcare utilisation. The primary comparative outcome is Days Alive and Out of Hospital (DAOH) over 12 months. Health-economic analyses will compare healthcare utilisation and direct healthcare costs across secondary care, primary care, and prescribing, with social care outcomes explored where data completeness permits. Together, these components aim to determine whether MinderCare can accurately detect clinically relevant change, be implemented feasibly within routine care, and show potential to improve real-world outcomes for PLWD. Within the comparative-effectiveness component, we will follow a target trial emulation (TTE) framework by specifying the pragmatic trial that would ideally compare activated MinderCare plus usual care with usual care alone, then emulating its eligibility criteria, treatment strategies, time zero, follow-up, outcomes, causal contrast and analysis plan using Discover-NOW linked health and care data [37].

## (B) OBJECTIVES

- To evaluate the performance of MinderCare digital biomarkers in detecting clinically relevant events against clinical ground-truth data.
- To assess changes in participant and carer wellbeing, functioning, and quality of life during MinderCare follow-up.
- To assess the feasibility, acceptability, sustainability, and resource requirements of implementing MinderCare within routine NHS and social care pathways.
- To estimate the effect of activated MinderCare, compared with usual care, on Days Alive and Out of Hospital over 12 months using a target trial emulation with an electronically matched external control cohort.
- To estimate the effect of MinderCare, compared with matched controls, on healthcare utilisation, healthcare costs, institutionalisation, and mortality over 12 months.

## (C) MINDERCARE OVERVIEW

This study is a mixed-methods protocol for the evaluation of MinderCare, a digitally enabled dementia care model that uses passive in-home sensors and clinical review to detect clinically relevant change and support timely intervention. The protocol comprises four linked components: (a) the digitally enabled remote monitoring care pathway; (b) the prospective evaluation of digital biomarker performance and implementation; (c) the comparative effectiveness using a target trial emulation with an electronically matched external control cohort; and (d) health economic analysis (Figure 1). MinderCare is sponsored by the UK Dementia Research Institute at Imperial College London in partnership with Imperial College Healthcare NHS Trust.

**Figure 1.**
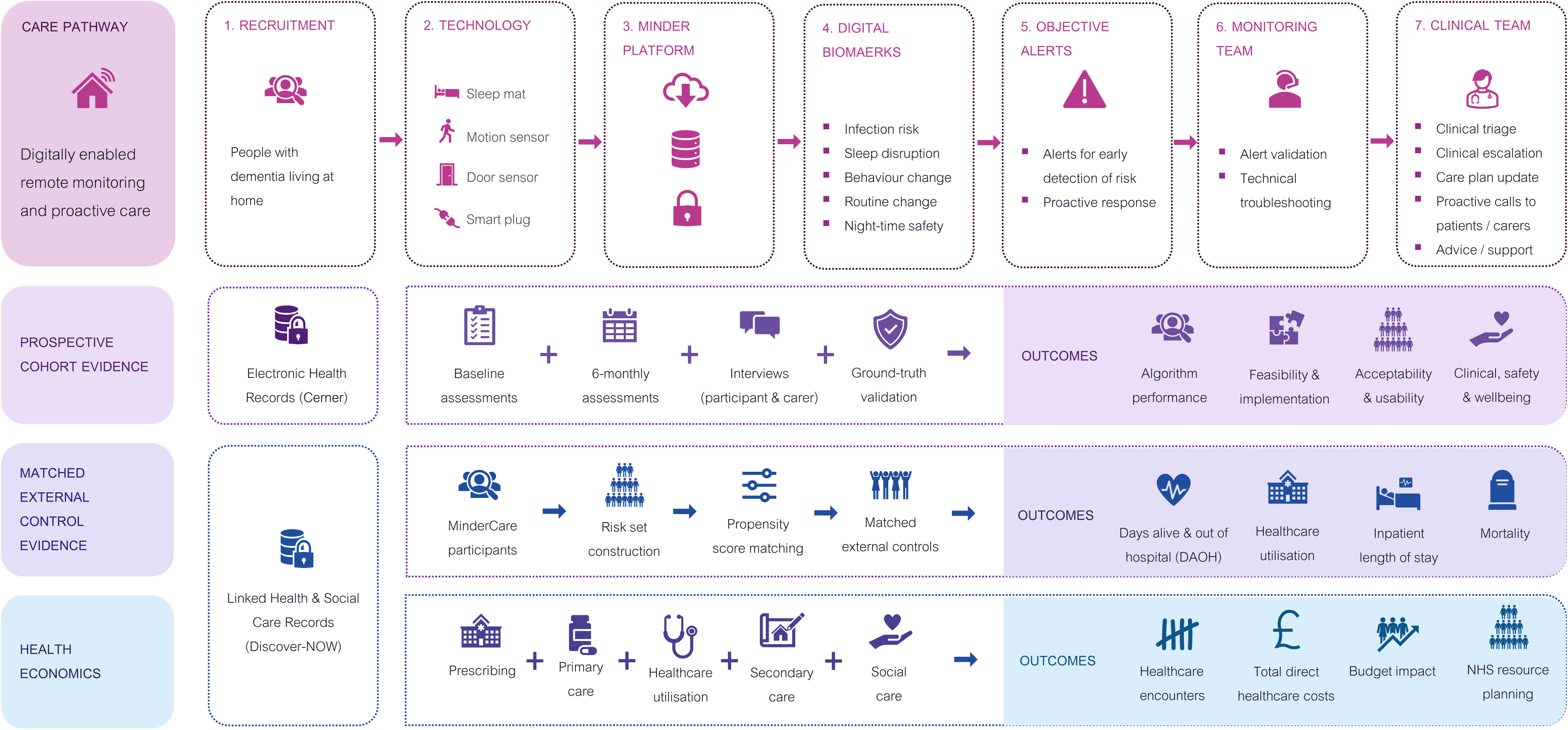
MinderCare study overview. The figure summarises the four components of the MinderCare protocol: A) Schematic of the digitally enabled remote monitoring care pathway and integrated evaluation framework. The three components of the evaluation include; B) prospective evaluation of digital biomarker performance and implementation, including feasibility, acceptability, and safety; C) comparative effectiveness using a target trial emulation with an electronically matched external control cohort derived from linked health and social care records; and D) health economic analysis of healthcare utilisation, costs, and system impact.

## (D) CARE PATHWAY

### (a) Study design

The study is delivered within routine NHS and social care pathways in North West London. Sensor-derived information is reviewed by the MinderCare team and, where clinically relevant, shared with professionals involved in the participant’s care through approved NHS communication routes.

Participants are enrolled for an initial 6-month monitoring period. They may continue for up to 12 months unless they choose to withdraw earlier. After 12 months, participants have the option to extend their involvement if they wish. Written informed consent is obtained from participants who have capacity. For individuals who lack capacity to consent, advice is sought from a personal consultee or, where necessary, a nominated consultee in accordance with applicable UK guidance.

After enrolment, UKCA-marked passive sensors are installed in the participant’s home (see Devices section). Baseline assessments are completed at or around installation by the PLWD and, where available, their study partner. These assessments include standardised measures of clinical status, wellbeing, functioning, carer experience, and service use. Follow-up assessments are repeated at 6 months and again at withdrawal or study exit (Table 1).

**Table 1.**
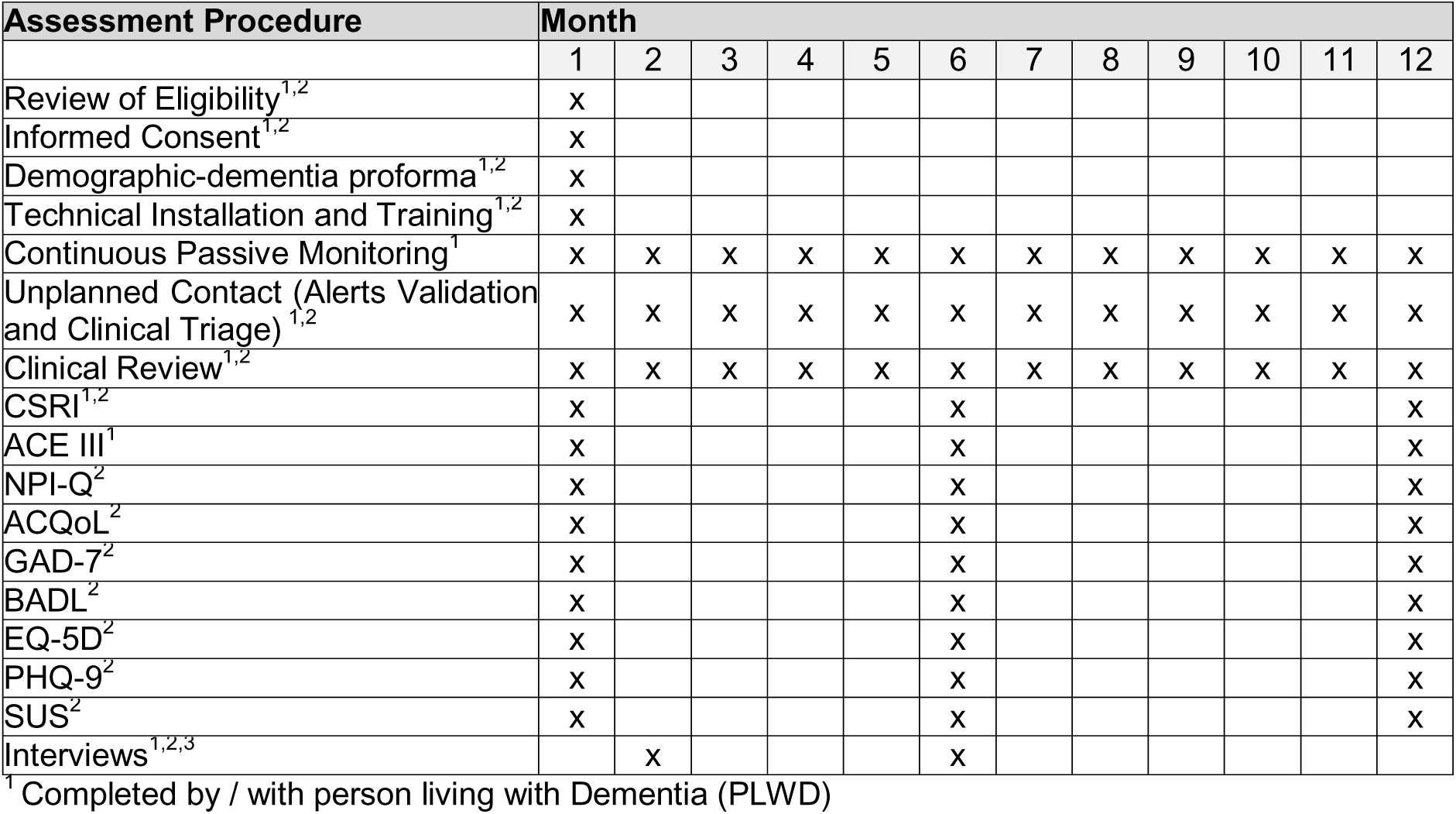
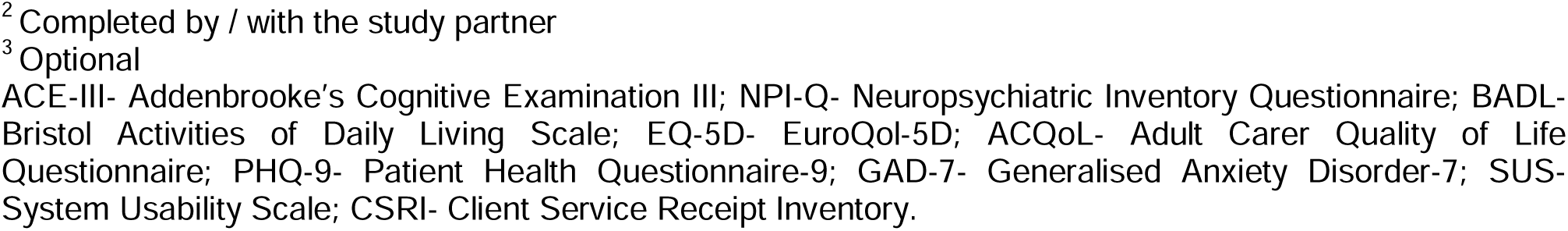
MinderCare Prospective Cohort: timeline and assessments.

| Assessment Procedure | Month |  |  |  |  |  |  |  |  |  |  |  |
| --- | --- | --- | --- | --- | --- | --- | --- | --- | --- | --- | --- | --- |
|  | 1 | 2 | 3 | 4 | 5 | 6 | 7 | 8 | 9 | 10 | 11 | 12 |
| Review of Eligibility<sup>1,2</sup> | x |  |  |  |  |  |  |  |  |  |  |  |
| Informed Consent<sup>1,2</sup> | x |  |  |  |  |  |  |  |  |  |  |  |
| Demographic-dementia proforma<sup>1,2</sup> | x |  |  |  |  |  |  |  |  |  |  |  |
| Technical Installation and Training<sup>1,2</sup> | x |  |  |  |  |  |  |  |  |  |  |  |
| Continuous Passive Monitoring<sup>1</sup> | x | x | x | x | x | x | x | x | x | x | x | x |
| Unplanned Contact (Alerts Validation and Clinical Triage)<sup>1,2</sup> | x | x | x | x | x | x | x | x | x | x | x | x |
| Clinical Review<sup>1,2</sup> | x | x | x | x | x | x | x | x | x | x | x | x |
| CSRI<sup>1,2</sup> | x |  |  |  |  | x |  |  |  |  |  | x |
| ACE III<sup>1</sup> | x |  |  |  |  | x |  |  |  |  |  | x |
| NPI-Q<sup>2</sup> | x |  |  |  |  | x |  |  |  |  |  | x |
| ACQoL<sup>2</sup> | x |  |  |  |  | x |  |  |  |  |  | x |
| GAD-7<sup>2</sup> | x |  |  |  |  | x |  |  |  |  |  | x |
| BADL<sup>2</sup> | x |  |  |  |  | x |  |  |  |  |  | x |
| EQ-5D<sup>2</sup> | x |  |  |  |  | x |  |  |  |  |  | x |
| PHQ-9<sup>2</sup> | x |  |  |  |  | x |  |  |  |  |  | x |
| SUS<sup>2</sup> | x |  |  |  |  | x |  |  |  |  |  | x |
| Interviews<sup>1,2,3</sup> |  | x |  |  |  | x |  |  |  |  |  |  |

<sup>1</sup> Completed by / with person living with Dementia (PLWD)
<sup>2</sup> Completed by / with the study partner
<sup>3</sup> Optional
ACE-III- Addenbrooke's Cognitive Examination III; NPI-Q- Neuropsychiatric Inventory Questionnaire; BADL- Bristol Activities of Daily Living Scale; EQ-5D- EuroQol-5D; ACQoL- Adult Carer Quality of Life Questionnaire; PHQ-9- Patient Health Questionnaire-9; GAD-7- Generalised Anxiety Disorder-7; SUS- System Usability Scale; CSRI- Client Service Receipt Inventory.

**Table 2.**
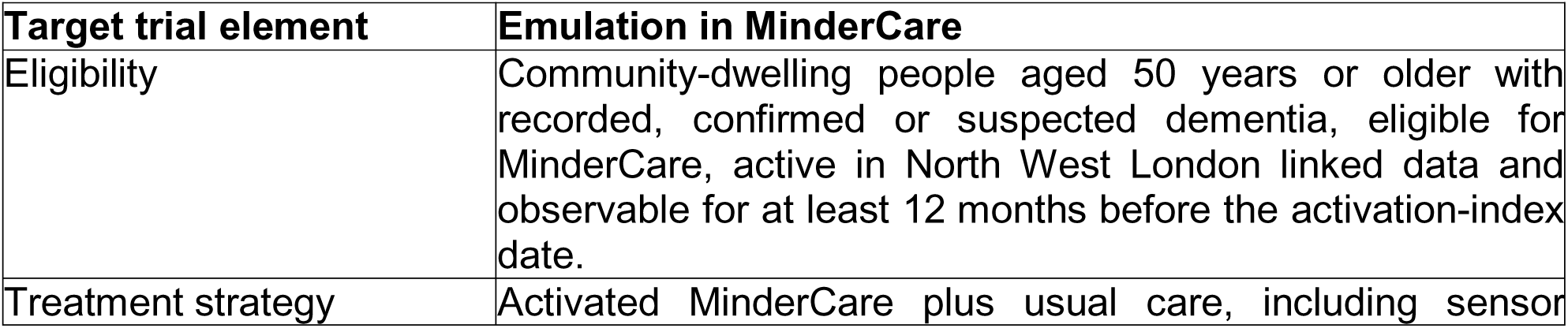
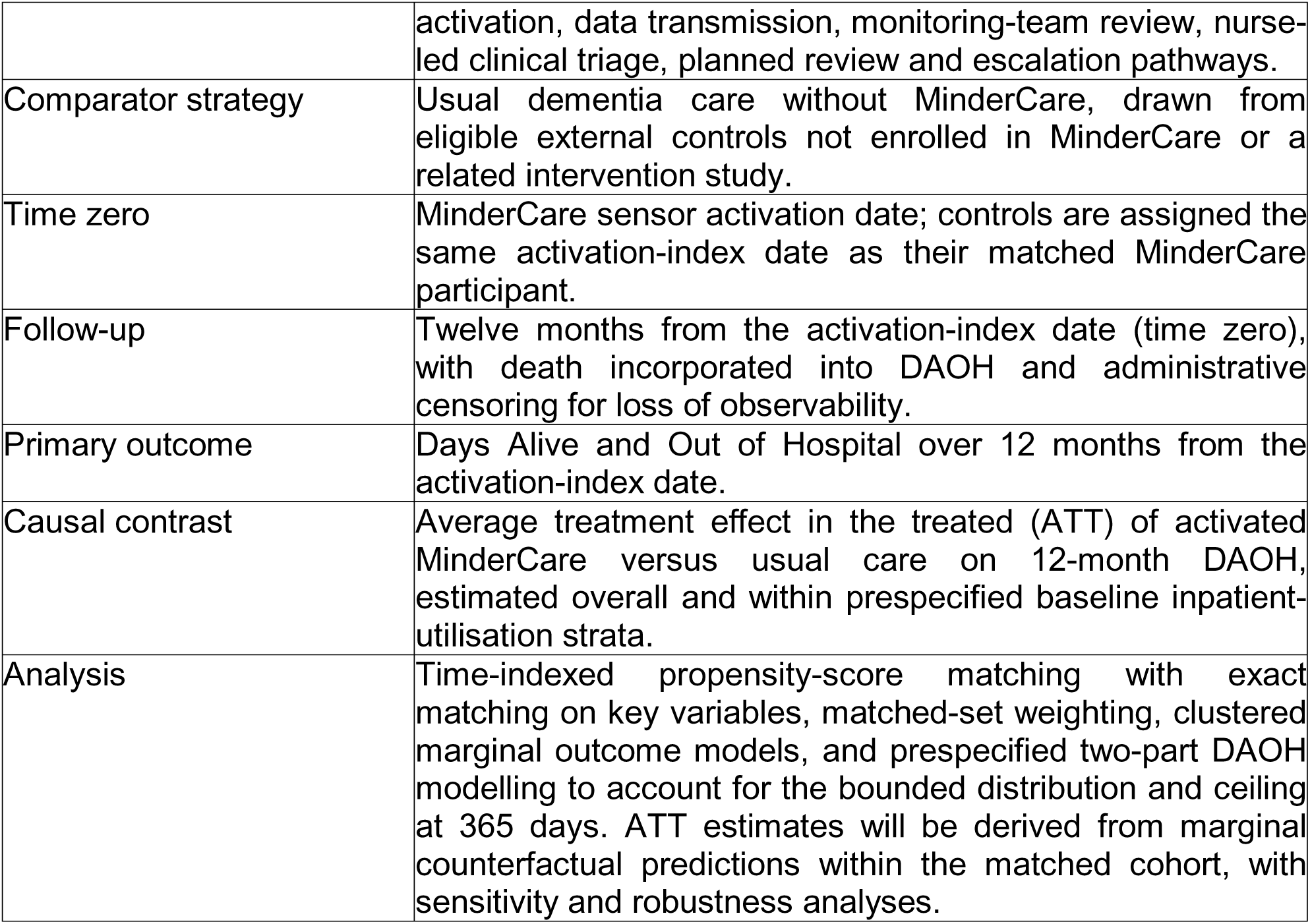
Target trial specification and emulation in MinderCare.

| **Target trial element** | **Emulation in MinderCare** |
| --- | --- |
| Eligibility | Community-dwelling people aged 50 years or older with recorded, confirmed or suspected dementia, eligible for MinderCare, active in North West London linked data and observable for at least 12 months before the activation-index date. |
| Treatment strategy | Activated MinderCare plus usual care, including sensor |
|  | activation, data transmission, monitoring-team review, nurse-led clinical triage, planned review and escalation pathways. |
| Comparator strategy | Usual dementia care without MinderCare, drawn from eligible external controls not enrolled in MinderCare or a related intervention study. |
| Time zero | MinderCare sensor activation date; controls are assigned the same activation-index date as their matched MinderCare participant. |
| Follow-up | Twelve months from the activation-index date (time zero), with death incorporated into DAOH and administrative censoring for loss of observability. |
| Primary outcome | Days Alive and Out of Hospital over 12 months from the activation-index date. |
| Causal contrast | Average treatment effect in the treated (ATT) of activated MinderCare versus usual care on 12-month DAOH, estimated overall and within prespecified baseline inpatient-utilisation strata. |
| Analysis | Time-indexed propensity-score matching with exact matching on key variables, matched-set weighting, clustered marginal outcome models, and prespecified two-part DAOH modelling to account for the bounded distribution and ceiling at 365 days. ATT estimates will be derived from marginal counterfactual predictions within the matched cohort, with sensitivity and robustness analyses. |

Sensor data collected from homes is sent to the Minder platform where the developed digital biomarker algorithms run at scheduled times and generate reports and alerts for the participants in the cohort. These are reviewed daily by the monitoring and clinical teams and are acted upon according to the relevant algorithm’s standard operating procedure. functions (Figure 2). The monitoring team undertakes first-line review of alerts, distinguishes likely technical artefacts from potentially clinically relevant changes, and escalates validated concerns. The monitoring team also supports device installation and technical troubleshooting. The clinical team reviews escalated alerts, undertakes clinical triage, contacts participants or carers as needed, updates care plans, provides advice, and liaises with relevant health and social care professionals.

**Figure 2.**
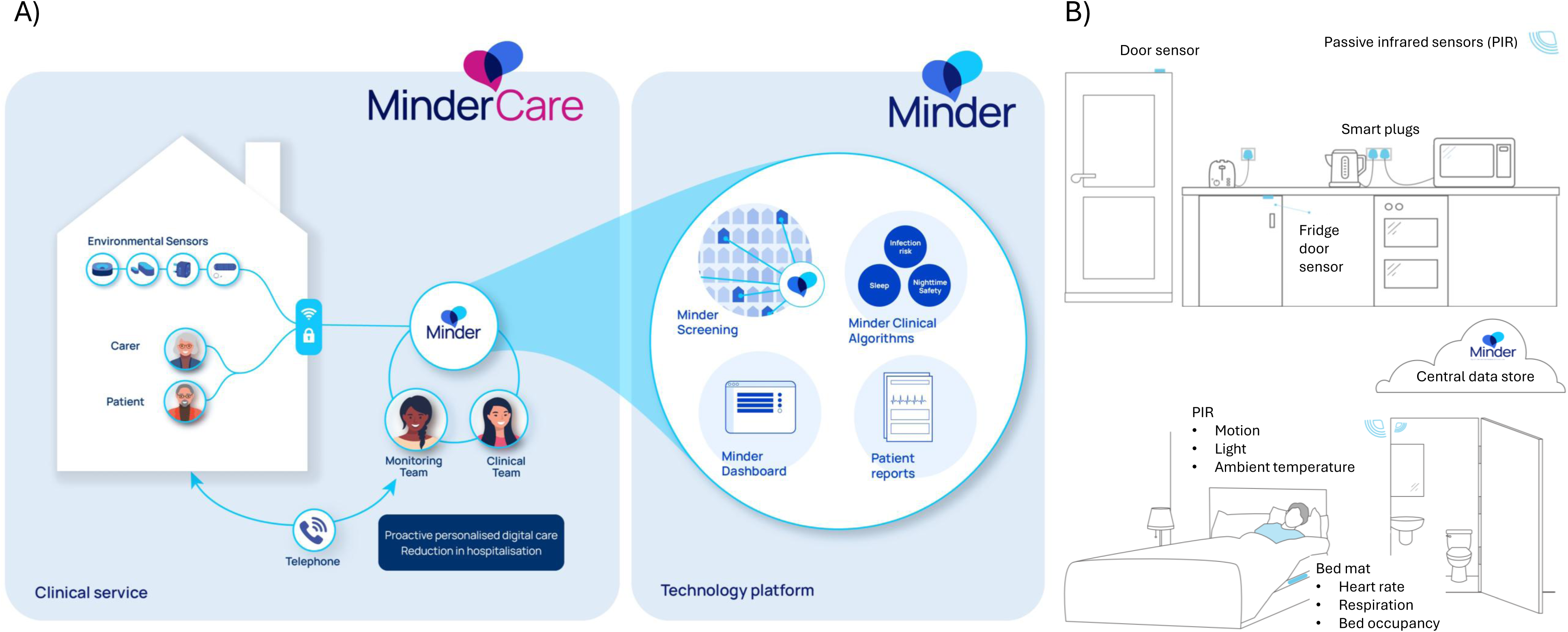
A) MinderCare workflow for the prospective cohort: Data generated by sensors installed in participants’ homes are collected through the Minder platform and reviewed by the monitoring and clinical teams to inform clinical decision-making. B) Deployment of devices in the homes of participants.

Participants and, where available, study partners are contacted monthly for planned clinical review and, on an ad hoc basis, in response to alerts or technical issues. These contacts support continuity of care, provide ground-truth information on symptoms, events, and care changes, and identify risks or needs not captured by sensors alone. Contacts may occur by telephone, video call, or email, according to participant preference and clinical need, and the nature, duration, and outcome of each contact are recorded.

### (b) Prospective cohort

We will recruit 100 people living with confirmed or suspected dementia into MinderCare. This target is pragmatic and is intended to support the four linked components of the protocol. For the digital biomarker component, a sample of this size is expected to provide sufficient alert-level and participant-level observations to estimate key performance metrics with 95% confidence intervals across a range of clinical and behavioural outcomes, informed by experience from the related Minder study (see relevant section below). The matched external-control component is justified separately in the dedicated sample-size subsection and the appendix.

### (c) Patient recruitment

#### (i) Inclusion criteria

Participants are eligible if they: (1) have a confirmed or suspected diagnosis of dementia of any subtype; (2) are aged 50 years or older at baseline; and (3) if they lack capacity to provide informed consent, have a personal or nominated consultee available. Where a study partner is involved, they must be aged 18 years or older and willing and able to provide informed consent. The absence of a study partner is not an exclusion criterion. The study is intended to include people who live alone or are socially isolated. Potential participants who do not fully understand spoken or written English may be included with translation or interpreter support, provided that consent or consultee procedures can be completed appropriately and study procedures can be delivered safely. Accessible participant materials, including visually led leaflets and brochures, are used to support understanding across a range of cognitive and language abilities. These materials were co-designed with PLWD through the Human-Centred Design group at Imperial College London.

#### (ii) Exclusion criteria

Participants are excluded if they: (1) have unstable comorbid mental illness requiring secondary mental health care at screening or baseline, including active psychosis or substance misuse; (2) have active suicidal ideation; (3) are receiving treatment for a terminal illness or are under palliative care at screening or baseline; or (4) lack capacity and do not have a personal or nominated consultee. Where a study partner is involved, they are excluded only if they are unable to participate in study procedures, including communication required for consent and follow-up, or unable to provide written informed consent.

#### (iii) Withdrawal criteria

In accordance with the Declaration of Helsinki, participants may withdraw from the study at any time, for any reason, without affecting their future medical care. Participants who withdraw stop all research-related activities. We ask withdrawing participants for consent to continue data linkage to their Electronic Health Records (EHRs) stored in the DISCOVER-NOW Database to monitor health and social care utilisation in relation to disease progression. Participants who decline ongoing data sharing are withdrawn completely, and no further data are collected.

#### (iv) Recruitment streams

Participants may be recruited through four routes: (1) clinical identification within Imperial College Healthcare NHS Trust, including community geriatric and dementia services; (2) referral from NHS and social care services, including GP practices, memory clinics, dementia link workers, community services, social services, assistive technology services, and careline teams; (3) third-sector organisations and self-referral; and (4) re-contact of individuals who have previously consented to be approached about research in other Imperial College studies. For routes involving prior contact by clinical or service teams, permission to share contact details with the MinderCare team is obtained before direct approach.

### (d) Devices

MinderCare uses passive sensors that do not require active operation by the participant or study partner. Sensor configuration is tailored to the home environment and participant preference and may include the following devices: (1) Sleep mat: A pressure-based sensor placed under the mattress (Withings Sleep Analyzer) that measures heart rate and respiration rate during sleep. (2) Movement sensors: Passive infrared (PIR) or radar-based sensors that detect presence or movement. (3) Smart plugs: Devices placed between an appliance plug and a mains socket to detect when appliances (e.g., kettles, microwaves) are used. (4) Door sensors: Sensors placed on the front door, back door, and fridge door to record when doors are opened or closed. (5) Gateway hub: A central hub that communicates with all environmental sensors (e.g., via the Zigbee protocol) and connects to the internet router. Participants without home Wi-Fi receive a study-provided router. Installation is undertaken in the participant’s home by the MinderCare monitoring team. The visit includes placement of sensors, confirmation of connectivity, explanation of device function, and an opportunity to address questions or concerns. Installation or re-installation usually takes less than 2 hours. Participants may opt for a reduced sensor configuration, including sleep-mat-only installation, if preferred.

### (e) Minder Clinical Algorithms (MCA)

#### (i) Overview

MinderCare uses a suite of sensor-derived clinical algorithms designed to identify patterns that may indicate clinically relevant change. These algorithms have been developed within the UK Dementia Research Institute Care Research and Technology programme and are deployed within MinderCare only after internal review, development of algorithm-specific standard operating procedures, and governance sign-off. During the initial phase of MinderCare, the following algorithms are implemented:

- Minder Infection Risk: detects sustained physiological and behavioural changes consistent with elevated infection risk [38], using sleep-derived respiratory and heart-rate measures together with contextual sensor data.
- Minder Sleep Index: identifies changes in sleep pattern and sleep disruption that may indicate worsening symptoms, distress, or emerging clinical problems [39,40].
- Activities and Routines: identifies clinically relevant change in activities of daily living and household routines, including reductions in activity, altered kitchen use, and potentially unsafe behaviours such as leaving doors open) [6].
- Night-Time Safety: uses door, movement, and sleep-sensor data to identify potential night-time safety risks, including nocturnal wandering and prolonged absence from bed or home.
- For each prespecified analysis, the deployed algorithm version, operating threshold and standard operating procedure will be documented. Analyses of subsequent model updates or retraining will be reported separately or treated as exploratory.

Algorithms will be iteratively developed and retrained as new data becomes available. In addition, new algorithms may be incorporated as these become available.

#### (ii) Reports & Alerts

Sensor-derived outputs are presented through the Minder dashboard as daily summaries and alert reports for the MinderCare monitoring and clinical teams. Relevant summaries may also be shared with GPs and other professionals involved in care through approved NHS communication channels, including the London Care Record and NHS mail, where appropriate to clinical management. Participants or study partners may request access to reports.

Alerts are generated when predefined algorithm thresholds or event rules are met. Alerts considered clinically relevant are discussed in daily multidisciplinary review with the clinical team, documented, and triaged to an action such as no further action, participant or carer contact, advice, care-plan update, or escalation to usual care services.

## (E) PROSPECTIVE COHORT EVIDENCE

### (a) Service delivery and implementation evaluation

#### (i) Questions

- What is the reach, retention, usability, technical performance, operational workload, and acceptability of MinderCare when delivered within routine NHS and social care pathways?
- What associations are observed between participant characteristics, carer wellbeing, and monitoring patterns such as alerts, clinical contacts, interventions, and workload?

#### (ii) Analysis Plan

The MinderCare clinical service will be evaluated using the RE-AIM framework [41,42] and the PRISM framework [43] to assess feasibility and sustainability. RE-AIM provides a structure for assessing reach and effectiveness. PRISM adds to the wider context of service delivery and implementation [41]. This dual approach evaluates how technical reliability and clinical workflow integration concurrently affect the feasibility of scaling the service across the Integrated Care System (ICS) [44]. Quantitative analyses will be primarily descriptive, summarising patterns of service reach, clinical need, effectiveness, user acceptance, operational delivery, and sustainability within the MinderCare model. Relationships between patient and service characteristics and implementation indicators will be examined using regression models selected according to the distribution and structure of the data. For count outcomes Poisson/negative binomial regression models will be used as appropriate, for numerical outcomes linear regression models, for binary outcomes logistic models and for categorical outcomes multinomial logistic regression models. These analyses are exploratory and intended to characterise real-world patterns of service delivery, clinical need, effectiveness, and user engagement within the MinderCare model. Interview and focus groups transcripts will undergo thematic analysis to explore user experience, acceptability, perceived reassurance, perceived continuity of care, and staff integration of monitoring information into clinical workflows. Retention at 12 months and operational indicators will be summarised descriptively to assess feasibility, acceptability, and longer term sustainability within the ICS.

#### (iii) Outcomes

##### (i) Clinical and well-being outcomes

- Cognitive function, neuropsychiatric symptoms, and activities of daily living, assessed using the Addenbrooke’s Cognitive Examination-III (ACE-III), Neuropsychiatric Inventory Questionnaire (NPI-Q), and Bristol Activities of Daily Living (BADL) Scale.
- Participant- and carer-reported quality of life, depression, and anxiety, measured using the EuroQol-5D (EQ-5D), Adult Carer Quality of Life questionnaire (ACQoL), Patient Health Questionnaire-9 (PHQ-9), and Generalised Anxiety Disorder-7 (GAD-7).

##### (ii) Healthcare utilisation

- Hospital admissions, GP consultations, care transitions, and mortality, derived from linked electronic health records and population health datasets (e.g. Discover NOW), and the Client Service Receipt Inventory (CSRI).

##### (iii) Implementation and experience

- Usability, user satisfaction, perceived ease of use, and intention to use, assessed via the System Usability Scale.
- Qualitative experience, including perceived usefulness, timeliness, and professional satisfaction, explored through semi-structured interviews and focus groups.
- System performance indicators, including Wi-Fi availability, device reliability, and reasons for non-enrolment, obtained from technical contact logs and qualitative data.

##### (iv) Safety

- Technical failures, adverse events, and system downtime, recorded through serious adverse event logs and system monitoring reports.

##### (v) Baseline contextual characteristics

- Participant and household characteristics, including language, ethnicity, social deprivation, and living situation, collected via a baseline dementia proforma.

### (b) Evaluation of MCAs

#### (i) Questions

- What is the alert-level and event-level performance of the Minder Clinical Algorithms for detecting clinically relevant events compared with clinical ground-truth data?
- How do algorithm performance metrics vary by event type, data completeness, participant characteristics, algorithm configurations, and context of use?

#### (ii) Analysis Plan

Minder Clinical Algorithms will be evaluated prospectively for real-world performance against clinical ground-truth data obtained through routine monitoring review, participant or carer contact, and relevant clinical records. The primary unit of analysis will be the alert, with event-level analyses undertaken where appropriate. For each algorithm, performance will be summarised using sensitivity, specificity, positive predictive value, negative predictive value, false-alert rate, and timeliness of detection. Estimates will be reported overall and, where feasible, stratified by algorithm type, event type, data completeness, and context of use. Repeated alerts within the same participant will be summarised descriptively, and participant-level summaries will be used where appropriate to avoid over-interpretation of multiple alerts arising from the same underlying event. This prospective validation framework has been implemented on the ground as part of standard operating procedures, with structured capture of whether alerts and observations generated from the digital biomarker algorithms correspond to clinically meaningful events.

#### (iii) Outcomes

##### (i) Performance of the algorithm against ground truth

- Sensitivity, Specificity, Positive predictive value, Negative predictive value, False-alert rate, Area under the ROC curve.

##### (ii) Usefulness of the algorithm

- Timeliness of detection, Clinical response ratio per response pathway, Time from alert to action

##### (iii) Practicality of the algorithm

- Estimated alert burden per 100 participants, Alert distribution per cohort and per participant, Data completeness and missingness rate, Redundant alert ratio.

##### (iv) Safety and equity of the algorithm

- Stratification of performance and alert coverage per age, sex, diagnosis, disease severity, and serious missed events rate (derived from false negatives that resulted in SAEs)

## (F) TARGET TRIAL EMULATION USING AN ELECTRONICALLY-MATCHED EXTERNAL CONTROL COHORT

### (a) Study design

The comparative-effectiveness component will be reported as a target trial emulation (TTE) nested within the wider mixed-methods protocol. It specifies the pragmatic trial that will compare activated MinderCare plus usual care with usual care alone, then emulates the key trial elements using linked electronic health record data from Discover-NOW (Figure 3). The emulated elements include eligibility criteria, treatment strategies, assignment and time zero, follow-up, outcomes, causal estimand and analysis plan. The intervention strategy comprises successful sensor installation and activation with data available for monitoring team and clinical review. The comparator strategy is usual care without MinderCare. The primary causal estimand is the average treatment effect in the treated (ATT) for participants under active monitoring, comparing their observed outcomes with the outcomes expected under usual care.

**Figure 3.**
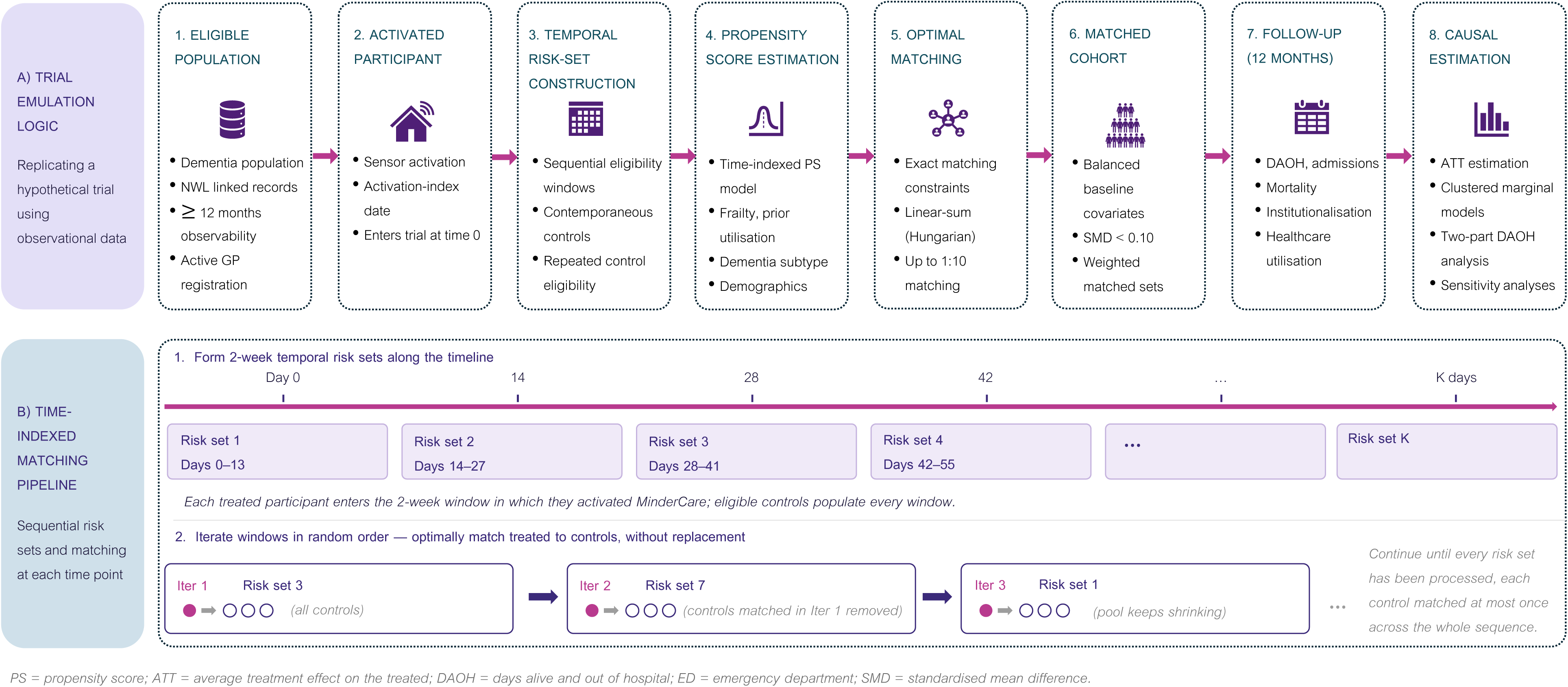
Sequential target trial emulation and time-indexed matching pipeline for the MinderCare comparative-effectiveness analysis. A) The upper panel summarises the emulated trial sequence: identification of an eligible North West London linked-record population with coded dementia or prespecified evidence of cognitive impairment/suspected dementia; MinderCare sensor activation defining time zero; construction of contemporaneous 14-day temporal risk sets; propensity-score estimation using baseline demographic, clinical, dementia-subtype, frailty, deprivation, and prior-utilisation variables; optimal matching with exact constraints; formation of weighted matched sets; 12-month outcome ascertainment; and ATT estimation for DAOH and secondary outcomes. B) The lower panel illustrates the sequential risk-set matching process. Each treated participant enters the 14-day window in which MinderCare is activated; eligible controls may contribute to multiple windows until selected; and matching is conducted iteratively without replacement until eligible risk sets have been processed. ATT, average treatment effect in the treated; DAOH, days alive and out of hospital; ED, emergency department; PS, propensity score; SMD, standardised mean difference.

### (b) Electronically-matched control construction

Controls are identified within the Discover-NOW dataset, a linked health and care resource covering more than 2.7 million individuals registered with GP practices across eight boroughs in North West London (Brent, Ealing, Hammersmith and Fulham, Harrow, Hillingdon, Hounslow, Kensington and Chelsea, and Westminster). The dataset integrates coded information from primary care, acute hospitals, mental health services, community services, and social care, enabling longitudinal tracking of patient pathways across the regional ICS.

Eligible individuals must be aged 50 years or older, have a recorded diagnosis of dementia or evidence of cognitive concerns on or before the assigned index date, be actively registered with a North West London GP practice, be community-dwelling, and have sufficient linked data for baseline covariate derivation. For each MinderCare participant, the index date will be the sensor activation date, defined as the date on which the home sensor system is activated and begins transmitting data for monitoring-team and clinical review. Matched controls will be assigned the same activation-index date as their matched MinderCare participant and must be alive, observable in Discover-NOW, actively registered, community-dwelling, and otherwise eligible on that date.

Eligible MinderCare participants and candidate controls must have at least 12 months of linked data before the activation-index date and at least one recorded GP contact during that baseline period. Individuals will be excluded if they died, transferred out, or were de-registered before index; were resident in long-term institutional care at index; were enrolled in MinderCare or another related intervention study; lacked sufficient baseline observability; or had incomplete data on the final matching variables.

### (c) Propensity score matching

Each MinderCare participant will be matched to up to 10 controls using a sequential, time-indexed propensity-score matching approach [45,46]. A person-period risk-set dataset will first be constructed across all 14-day eligibility buckets. Candidate controls may contribute to more than one bucket while they remain eligible, reflecting the fact that they could have been selected as a comparator at different calendar times [37]. Propensity scores will then be estimated across this full time-indexed dataset using logistic regression, with treatment status defined by MinderCare participation and baseline covariates measured before the relevant activation-index date.

Matching will be performed within each 14-day eligibility bucket, so that MinderCare participants are compared only with controls who were eligible and observable in the same temporal risk set. Within each bucket, controls will be selected using optimal linear-sum assignment [47,48] subject to a caliper of 0.2 standard deviations of the logit of the propensity score [49], exact matching constraints, and the target of up to 10 controls per treated participant. Buckets will be processed iteratively in randomised order to reduce potential order effects when controls are matched without replacement.

The prespecified baseline features for propensity-score estimation will include age at the matching time point, age band, sex, ethnicity category, Index of Multiple Deprivation quintile, local authority district of residence, dementia subtype, baseline frailty summary [50], and prior healthcare utilisation. Prior healthcare utilisation will be defined using total inpatient bed-days in the 12 months before the activation-index date: 0, 1–9, 10–29, and ≥30 bed-days. Exact matching will be applied on age band, sex, local authority district of residence, and prior healthcare utilisation category. Covariate balance will be assessed using absolute standardised mean differences, with values below 0.10 taken to indicate acceptable balance. Where clinically important residual imbalance remains after matching, the imbalanced covariates will be included as adjustment terms in outcome models.

### (d) Sample size for cohort and matched control arm

To determine the statistical power for the comparison between MinderCare and the electronically matched controls, we conducted a simulation study. This involved generating hypothetical trial data based on real-world data from individuals aged ≥50 years with dementia drawn from the Discover-NOW dataset, allowing us to estimate the likelihood of detecting a true treatment effect. Full details of the simulation methods and underlying assumptions are provided in the appendix. Under a scenario with 100 MinderCare participants and five matched controls per participant (5:1 matching ratio), yielding a total analytic sample of 600 individuals (100 intervention, 500 controls), the study is estimated to have 88% power to detect a 15% improvement in Days Alive and Out of Hospital (DAOH) over 12 months at a two-sided significance level of 0.05. Under the same assumptions, the study has 79% power to detect a 12% improvement in DAOH. In absolute terms. These effect sizes correspond to approximately an average of 5.1–5.7 additional DAOH over 12 months for MinderCare compared to controls.

### (e) Evaluation of MinderCare effectiveness

#### (i) Questions

- What is the average treatment effect in the treated of MinderCare on Days Alive and Out of Hospital (DAOH) over 12 months (see below for definition), compared with electronically matched external controls?
- How does participation in MinderCare relate to healthcare utilisation, institutionalisation, mortality, and other routine-care outcomes compared with matched controls?

#### (ii) Analysis Plan

##### (i) Estimand and analysis population

The primary estimand is the average treatment effect in the treated [45,46] (ATT) for Days Alive and Out of Hospital (DAOH) over 12 months following the activation-index date for MinderCare participants compared with usual care. The analysis will be conducted within a sequential target trial emulation framework [37] using an electronically matched external control cohort derived from the Discover-NOW [51] linked health and care dataset. For MinderCare participants, the activation-index date is defined as the date on which the home sensor system is activated and begins transmitting usable data for monitoring-team and clinical review. For candidate controls, eligibility will be assessed within the same 14-day temporal eligibility bucket as the corresponding MinderCare participant. Controls must be alive, observable, actively registered with a North West London GP practice, community-dwelling, and otherwise eligible at the relevant time point. The control index date will be assigned from the qualifying GP/contact date within the relevant temporal eligibility bucket, thereby establishing contemporaneous observability. The analysis population will comprise MinderCare participants and matched controls with sufficient linked data for baseline covariate derivation and follow-up outcome ascertainment. Within variable-ratio matched sets, controls will be weighted so that the total control weight within each matched set equals 1, giving each treated participant equal contribution to the ATT.

##### (ii) Two-part Days Alive and Out of Hospital sensitivity analysis

Because DAOH is bounded and may show substantial clustering at the upper boundary of 365 days, a prespecified two-part sensitivity analysis will be undertaken. First, a logistic model will estimate the probability of attaining the ceiling value of DAOH = 365 days. Second, among participants with DAOH below 365 days, a Gaussian model will estimate mean DAOH conditional on not achieving the ceiling value. Counterfactual predictions under MinderCare and usual care will then be combined to estimate marginal mean DAOH under each condition, the corresponding ATT, and the treatment effect on the probability of achieving the ceiling value.

##### (iii) Interpretation of the average treatment effect

The ATT represents the mean difference in 12-month DAOH between MinderCare participants and the outcome they would be expected to experience under usual care, conditional on the sequential target trial emulation framework, the matched external control design, measured baseline covariates, and the specified outcome model. As this is not a randomised trial, estimates will be interpreted as observational comparative-effectiveness estimates. Residual confounding from unmeasured or incompletely measured factors may remain despite time-indexed eligibility, propensity-score matching, exact matching constraints, and post-matching balance assessment.

##### (iv) Secondary comparative outcomes

Secondary comparative outcomes will be analysed using the matched cohort and models appropriate to the scale and distribution of each outcome. Analyses will account for matched-set clustering and matching weights where applicable. Healthcare utilisation outcomes, including emergency department attendances, hospital admissions, inpatient bed-days, outpatient appointments, and GP contacts, will be analysed using count or other suitable marginal models according to the observed distribution. Binary outcomes will be analysed using clustered logistic regression. Time-to-event outcomes, including mortality and institutionalisation where available, will be analysed using index-aligned survival models. State transitions among home, hospital, care home, and death will be summarised descriptively and, where feasible, modelled using multistate methods. Prespecified participant characteristics, including frailty and prior healthcare utilisation, will be summarised descriptively and incorporated into adjusted secondary analyses where relevant.

##### (v) Exploratory within-group comparisons

As exploratory descriptive analyses, outcomes measured over the 12-month follow-up period will also be compared with outcomes measured over the previous 12 months separately within the MinderCare group and within the matched control group. These within-group comparisons will be used to describe temporal change within each group and will not be interpreted as estimates of the comparative treatment effect. They will be analysed using paired or repeated-measures methods appropriate to the scale and distribution of each outcome.

##### (vi) Missing data and observability

Matching requires complete data on the final matching variables. Candidate controls with missing values in any matching variable will therefore be excluded from the matched control pool. MinderCare participants included in the comparative analysis must have sufficient linked data available for baseline covariate derivation and follow-up outcome ascertainment. No routine imputation of outcome data will be undertaken. For secondary time-based outcomes, later loss of observability due to deregistration, transfer out, or data cut will result in administrative censoring. For DAOH, sensitivity analyses will examine the impact of assumptions regarding the recorded month of death and post-index loss of observability.

##### (vii) Sensitivity and robustness analyses

Sensitivity analyses will include alternative matching ratios, for example, up to 1:5 and up to 1:3; alternative propensity-score specifications; alternative caliper or common-support restrictions; additional adjustment for residual post-matching imbalance; stability checks across different random seeds for bucket ordering; alternative assumptions for recorded month of death; and inverse-probability-of-censoring weighting if later loss of observability is material. The two-part DAOH model described above will be included as a prespecified sensitivity analysis. Where appropriate, E-values or similar methods may be used to explore the potential impact of unmeasured confounding on the primary estimate.

##### (viii) Reporting

We will report the number of MinderCare participants included, the number successfully matched, the distribution of matched-set sizes, the number of eligible controls available across temporal buckets, caliper or common-support exclusions, pre- and post-matching covariate balance, achieved propensity-score distances, and a study flow diagram describing control pool construction, exclusions, risk-set construction, and matching yield. Primary and secondary comparative estimates will be reported with confidence intervals, together with descriptive summaries of healthcare utilisation, DAOH, mortality, institutionalisation, and costs.

#### (iii) Outcomes

##### (i) Primary Outcome

The primary outcome for the comparative-effectiveness analysis, conducted within a sequential target trial emulation framework, is Days Alive and Out of Hospital (DAOH) over 12 months following the activation-index date. For MinderCare participants, the activation-index date is defined as the date on which the home sensor system is activated and begins transmitting usable data for monitoring-team and clinical review. Matched controls will be selected from the same 14-day temporal eligibility bucket as the corresponding MinderCare participant, with the control index date assigned from the qualifying GP/contact date within that bucket. DAOH is defined as the number of calendar days during the 12-month follow-up period in which a participant is alive and not admitted to hospital. DAOH is calculated according to the following rules:

- Death: Days following death contribute zero to DAOH (death is treated as part of the outcome process rather than as a censoring event). Due to limitations in data accuracy, the date of death will be assumed to be the first day of the recorded calendar month, with sensitivity analyses exploring alternative assumptions.
- Hospitalisation: Days overlapping an inpatient hospital spell are not counted as days out of hospital.
- Emergency department or outpatient visits: These are counted as days out of hospital unless they convert to an inpatient admission or cross midnight as an inpatient stay.
- Transfer out or deregistration: Follow-up contributes observable time until loss of data availability. The remainder of the window is administratively censored and DAOH is calculated over the observed portion.
- Maximum value: The maximum possible DAOH corresponds to the total number of days within the 12-month follow-up window (365 days).
- DAOH will also be summarised within prespecified strata of previous inpatient healthcare utilisation, defined using total inpatient bed-days in the 12 months before the activation-index date: 0 bed-days, 1-9 bed-days, 10-29 bed-days, and 30 or more bed-days. These categories distinguish participants with no recent inpatient use from those with low, moderate and high baseline hospital burden. Stratified summaries will be used to describe whether DAOH patterns and estimated MinderCare effects vary according to pre-index inpatient utilisation.

##### (ii) Secondary outcomes

The comparative component will be measured over the 12-month follow-up period unless otherwise specified. These include:

- Healthcare utilisation

◦ GP consultations
◦ Emergency department attendances
◦ Hospital admissions
◦ Inpatient bed-days
◦ Outpatient appointments
- Social care utilisation

◦ Measures such as domiciliary care use (where data are available and of sufficient quality)
- Clinical events

◦ Cause-specific hospitalisations (e.g., infections, trauma)
◦ Frequency and duration of hospital stays
- State transitions

◦ Movements between home, hospital, care home, and death during follow-up, analysed as a longitudinal multistate process
- Participant characteristics

◦ Prespecified factors such as frailty level (e.g., electronic frailty index) [50], wellbeing scores, and service use patterns, including ambulance calls and time to hospital readmission

## (G) PARTICIPANT AND STAKEHOLDER EXPERIENCE AND ENGAGEMENT

### (a) Evaluation of participant experience

To explore participant and carer experience, up to 20 participants and/or study partners will be invited to take part in optional semi-structured interviews at approximately 2 months after installation and again at 6 months or at de-installation (exit interview). Interviews will explore perceived usefulness, burden, acceptability, barriers and facilitators to use, and suggestions for improvement. Interviews may be conducted by telephone, video call, or home visit and will be analysed thematically.

### (b) Healthcare professionals engagement

Up to 12 health and social care professionals involved in MinderCare or in receiving its outputs will be invited to participate in surveys, focus groups, and follow-up interviews. These activities will inform co-design and refinement of clinical summary reports and will explore usability, clinical relevance, workflow fit, and implementation barriers and facilitators.

### (c) Patient and public involvement

Patient and public involvement and engagement (PPIE) has informed MinderCare from the outset. A dedicated Dementia Lived Experience Group (DLEG) of up to seven carers of PLWD contributes through regular meetings throughout study design and delivery. The group reviewed research questions, outcomes, participant-facing documents, questionnaires, and study equipment, and informed the development of recruitment materials, assessment of participant burden and perceived intrusiveness.

DLEG members have also reviewed and refined the 2-month, 6-month, and exit interview discussion guides, with a focus on making questions accessible, non-technical and non-judgemental. The questions have been designed to follow best-practice guidance informed by the group: one idea per sentence; short, clear questions; no “test” questions; supportive and conversational tone; avoidance of complex choices; and use of familiar, everyday words. DLEG input also supported ethical and practical decisions about how to conduct interviews in people’s homes. This included advice on handling potentially sensitive topics, such as health changes or hospital visits. Their contribution helped ensure that the interview is accessible, respectful, and supportive for participants and study partners, and aligned with the wider values of the MinderCare programme. PPIE contributors will also advise on dissemination, including which findings to share, with whom, and in what format. PPIE contributors are not enrolled as study participants.

## (H) HEALTH-ECONOMIC EVALUATION

### (a) Question

What are the differences in healthcare resource use and costs between MinderCare participants and matched controls, and what is the projected budget impact of wider implementation?

### (b) Analysis plan

This budget impact analysis (BIA) will follow established good practice guidance, including recommendations from the ISPOR. The BIA analysis is designed to estimate how introducing MinderCare would affect healthcare spending over the short to medium term, focusing on affordability and the likely changes to annual budgets rather than cost-effectiveness. The analysis will be conducted from the perspective of the NHS and Personal Social Services. Only relevant direct healthcare costs will be included. The time horizon will reflect typical budgeting cycles, covering a period of one to five years. The size of the population eligible for the intervention will be estimated using the best available epidemiological evidence alongside assumptions about diagnosis and treatment rates in routine practice. Where appropriate, changes in the population over time, such as growth or shifts in demographics, may also be taken into account.

Two scenarios will be compared. The first represents current practice without the new intervention, while the second reflects a situation in which the intervention is introduced. For each scenario, treatment pathways will be described in a way that captures how patients are actually managed in practice. The expected uptake of the new intervention will be modelled gradually over time, based on realistic assumptions informed by published evidence, experience with similar interventions, or expert input.

The analysis will include all relevant cost components. These are expected to cover the cost of the intervention itself (including acquisition and administration), as well as broader healthcare resource use such as clinic visits, hospitalisations, monitoring, and the management of adverse events. Unit costs will be drawn from appropriate and credible sources and reported using a clearly defined price year and currency. In line with standard practice for BIAs, costs will generally not be discounted.

Resource use and costs will be combined to estimate total spending in each scenario. These totals will then be compared to calculate the overall budget impact, expressed as the difference in annual costs between the scenario with MinderCare and the current standard of care. Results will be presented in a way that clearly shows both the total and incremental financial consequences over the analysis period. Given the uncertainty inherent in some inputs, sensitivity and scenario analyses will be carried out to test how results change when key assumptions vary. This will include varying parameters such as uptake rates, costs, and population estimates, as well as exploring alternative plausible scenarios. All key assumptions will be clearly stated and justified to ensure transparency. Finally, steps will be taken to ensure the model is robust and credible. This will include internal checks to confirm that calculations are consistent and, where possible, validation against published evidence or expert clinical opinion. The model will be implemented using transparent and accessible tools (e.g. Microsoft Excel, R, etc.).

### (c) Outcomes

Health-economic analysis will be performed using linked electronic health records and population health datasets to estimate healthcare resource use and associated costs for MinderCare participants compared with electronically matched controls. Outcomes will include:

#### (i) Secondary care costs

- Hospital admissions
- Inpatient bed-days
- Emergency department attendances
- Outpatient visits

#### (ii) Primary care costs

- GP clinical encounters
- Administrative encounters

#### (iii) Medication costs

- Medication costs derived from prescribing records

#### (iv) Total healthcare costs

- Aggregated direct healthcare costs across primary care, secondary care, and medications

#### (v) Social care costs

- Social care utilisation costs e.g. domiciliary care use (where data are available and of sufficient quality)

These outcomes will inform the health-economic evaluation and budget impact analysis assessing the potential financial implications of implementing MinderCare within NHS and social care systems.

## (I) ETHICS AND DISSEMINATION

MinderCare is funded by LifeArc and the UK Dementia Research Institute at Imperial College London. It is sponsored by Imperial College London in partnership with Imperial College Healthcare NHS Trust. MinderCare has received ethical approval from the North East – Newcastle and North Tyneside 2 Research Ethics Committee (IRAS 345123, REC reference 24NE0190) and has received confirmation of capacity and capability by the Imperial College Healthcare NHS Trust R&D. MinderCare is registered on the ISRCTN registry (ISRCTN14997677) and on the NIHR portfolio (CPMS ID 63023).

Participants (patients and study partners) provide written informed consent prior to the commencement of any study procedures. All participants are fully informed about the purpose of the study and the collection, use, storage, and handling of their data. If a patient lacks capacity to provide consent, a consultee – either personal (e.g., a carer) or professional (e.g., a GP) – will be consulted. For qualitative interviews with participants and focus groups with healthcare professionals, dedicated Participant Information Sheets and Consent Forms are provided.

Data is stored on Imperial College London–hosted servers within the secure University research cloud environment and in REDCap. Identifiable information is stored separately on an NHS secure drive, accessible only to authorised ICHT staff, with any hard-copy documents kept in a locked cabinet at the NHS site.

Data collected as part of the MinderCare study will be made available through the Dementia Platform UK in accordance with data-sharing policies. Study findings will be disseminated through presentations at national and international conferences, publication in peer-reviewed scientific journals and stakeholder engagement activities.

## Data Availability

All data produced in the present study are available upon reasonable request to the authors

## AUTHOR’S CONTRIBUTIONS

DJS, SD and ES conceived the study. JJ-J, MG, SD, MH, MT, SC, ENE, ES and DJS contributed to the development of the methodology. JJ-J, MT, SD, SC, HL, NJ, GF, MT, TS, ENE, ES and DJS drafted the manuscript. JJ-J and MT made equal first author contributions. ES and DJS made equal senior author contributions. DJS is the guarantor. All authors contributed to the design of the protocol and approved the final manuscript.

## ACKNOWLEDGMENTS

We thank the MinderCare Dementia Lived Experience Group for their valuable contributions throughout the project, which have helped ensure that its development remains aligned with the needs and experiences of people living with dementia. We also acknowledge the ongoing input of the CR&T group and thank Dr Aglaja Dar, Dr Pandora Wright, and Sarah Pearse for their guidance in shaping the delivery of the MinderCare clinical service.

## APPENDIX

### (a) Additional sample size simulation methods

Power for the electronically matched control arm comparison was explored using simulation. As this study is not designed to provide a definitive assessment of effectiveness, these analyses are intended to characterise the range of effect sizes that could be detected with the available MinderCare sample size, rather than to formally power a hypothesis test.

Simulations were used to examine the ability to detect absolute improvements of 12 or 15% in DAOH for MinderCare compared to electronically matched control patients for various different numbers of matched controls (1 to 10) and 80 to 100 MinderCare patients. These analyses are intended to inform interpretation of observed effects and to support the design and sample size calculation of a future definitive trial.

For each sample size explored, data on DAOH were initially simulated for individuals in the control.

Three potential patient pathways were considered during a 12 month follow-up:

- Died
- Did not die - with hospitalisation
- Did not die - no hospitalisation

Data from an external cohort of individuals aged 50 years and over, with all types of dementia, from the DiscoverNOW Databank in the year 2024 were used to inform the proportions of individuals expected in each of the above 3 patient pathways, and for each pathway, the mean and SD of DAOH. These were as follows:

**Table A1:** Summary statistics assumed for control patients in sample size simulations. For DAOH data presented as mean +/−SD.

| hospital_in_fu | died_in_fu | n | DAOH |
| --- | --- | --- | --- |
| False | False | 28713 | 365.91 +/- 0.29 |
| True | False | 7958 | 348.24 +/- 23.43 |
|  | True | 3769 | 141.10 +/- 105.13 |

DAOH for the control group were simulated for each patient pathway using Normal distributions for the two profiles who did not die and a Poisson distribution for those who died, with means as in Table A1. For the MinderCare group, the anticipated means for DAOH (days) for the active group were determined by inflating the control means by 12 or 15% for each of the three patient pathways, and data simulated using Normal or Poisson distributions with the inflated means. Improvements of 12% and 15% in the DAOH corresponded to an average absolute increase of around 5.1 and 5.7 days in DAOH for MinderCare relative to control. The maximum DAOH was set to be 365 days, given the 12 month follow-up period. The resulting data are bimodal with a spike at 365 days, as seen with this type of data.

For each simulated dataset, due to the distribution of DAOH (bi-modal with a peak at 365 days), a non-parametric Wilcoxon rank sum test was conservatively applied, and p-value extracted. A total of 1,000 simulations were conducted. Power was computed as the proportion of times a treatment effect was identified as statistically significant in this setup, where the true effect ranged from 12% to 15% increase (mean difference of 5.1 to 5.7 DAOH), taking a two-sided type 1 error rate of 0.05 across the 1,000 simulations. Simulations were performed using Stata (version 17).

### (b) Additional sample size results

In the best-case scenario where data is obtainable from 100 MinderCare patients, with 5 matched controls per MinderCare patient (5 controls:1 MinderCare) a total sample size of 600 (500 controls: 100 MinderCare) will provide 88% power to detect a 15% improvement (mean difference of 5.7 DAOH), or 79% power to detect a 12% improvement in DAOH (mean difference of 5.1 DAOH) over 12 months at a 5% significance level. If data are obtainable from 80 MinderCare patients, with 5 matched controls per MinderCare patient (5 controls:1 MinderCare), a total sample size of 480 (400 controls: 80 MinderCare) will provide 79% power to detect a 15% improvement (mean treatment group difference of 5.7 DAOH), or 65.8% power to detect a 12% improvement in DAOH over 12 months at a 5% significance level. Where data is obtainable from 90 MinderCare patients, with 5 matched controls per MinderCare patient (5 controls:1 MinderCare) a total sample size of 540 (450 controls: 90 MinderCare) will provide 84% power to detect a 15% improvement, or 75% power to detect a 12% improvement in DAOH over 10 months at a 5% significance level.

Power with a different number of matched controls to detect a 12% or 15% improvement in DAOH can be seen in Figures A1-A3.

**Figure A1:**
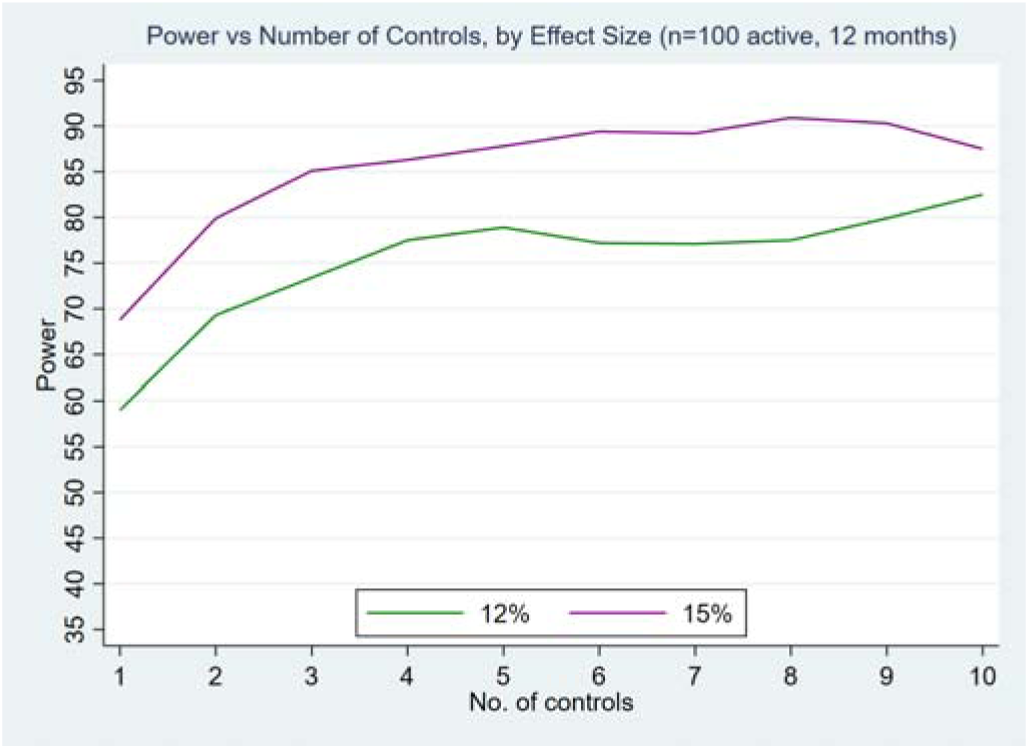
Power with n=100 MinderCare and 1 to 10 matched controls. Effect size is the improvement in DAOH for MinderCare relative to placebo.

**Figure A2:**
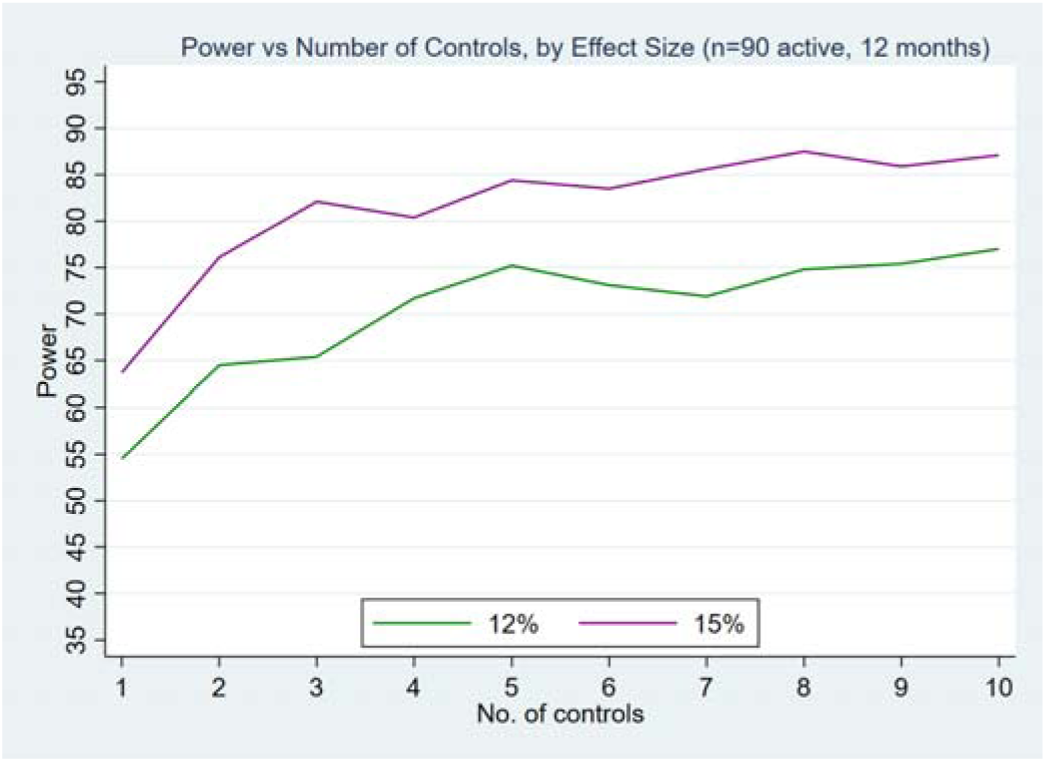
Power with n=90 MinderCare and 1 to 10 matched controls. Effect size is the improvement in DAOH for MinderCare relative to placebo.

**Figure A3:**
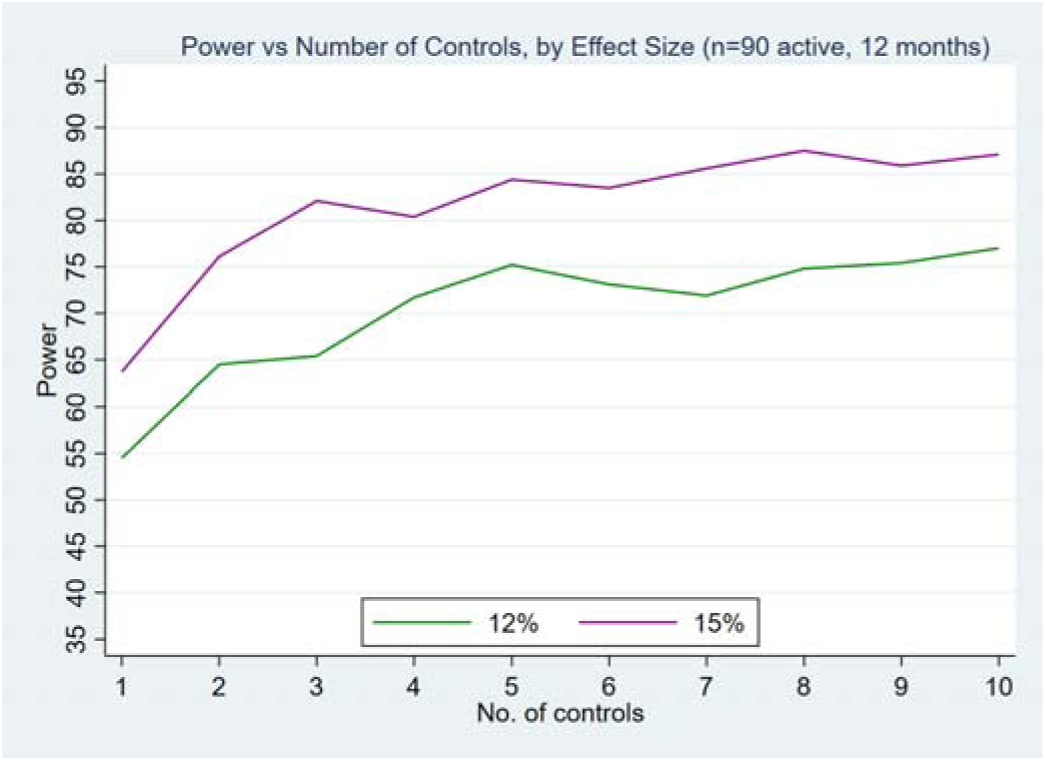
Power with n=80 MinderCare and 1 to 10 matched controls. Effect size is the improvement in DAOH for MinderCare relative to placebo.

### (c) Electronic matched control pipeline and data processing workflow

This appendix describes the data processing and matching pipeline used to construct the electronically matched external control cohort and the final analysis dataset for comparative effectiveness and health-economic evaluation.

#### (i) Overview

The matched control cohort is derived from the Discover-NOW linked electronic health record dataset using a structured, multi-stage pipeline. The pipeline constructs a contemporaneous risk set, derives baseline covariates and retrospective utilisation measures, performs propensity score matching, and generates an analysis-ready dataset including healthcare utilisation and cost outcomes. Each stage of the pipeline is applied sequentially, with intermediate outputs persisted to ensure reproducibility and transparency.

##### (i) Step 1. Construction of the eligible cohort

An initial eligible cohort is constructed comprising:

- MinderCare participants (treated group), and
- a pool of candidate control individuals drawn from the same underlying population.

Core inclusion and exclusion criteria are applied at this stage, including:

- age ≥50 years,
- recorded dementia diagnosis,
- active registration with a North West London GP practice, and
- sufficient data availability for baseline covariate derivation.

This step defines the initial treated cohort and the corresponding pool of eligible controls.

##### (ii) Step 2. Persistence of eligibility outputs

The eligible cohort is persisted as a set of intermediate tables to enable reproducibility and downstream processing. These tables contain identifiers for treated participants and eligible controls, along with key eligibility flags and index date information.

##### (iii) Step 3. Derivation of retrospective eligible sample

A retrospective eligibility step is applied to ensure comparability in baseline measurement and follow-up:

- Index dates are assigned to controls to align with the recruitment date of matched treated participants.
- A fixed retrospective baseline window (12 months prior to index) is defined.
- Retrospective utilisation measures are derived, including prior Days Alive and Out of Hospital (DAOH) and related summaries of healthcare use.
- Individuals without sufficient observability in the baseline period are excluded.

This step produces the final cohort eligible for matching.

##### (iv) Step 4. Baseline covariate construction

Baseline covariates are constructed and harmonised into a wide, person-level dataset with one row per individual at the index date. These include:

- demographic variables (e.g. age, sex, ethnicity, local authority district of residence, deprivation),
- clinical variables (e.g. dementia subtype),
- frailty or summary health status measures, and
- prior utilisation measures (e.g. inpatient bed-days, service use).

Variables are cleaned, encoded, and grouped where appropriate (e.g. age bands, utilisation categories) to support stable matching and interpretation.

##### (v) Step 5. Propensity score estimation

A propensity score model is fitted using logistic regression, with treatment status (MinderCare participation) as the dependent variable and baseline covariates as predictors. The model is estimated within a time-indexed framework so that treated participants and candidate controls are evaluated contemporaneously at the same index date. The resulting propensity score represents the probability of receiving the intervention conditional on observed baseline characteristics.

##### (vi) Step 6. Propensity score matching

Matching is performed to construct comparable treated and control groups:

- Each treated participant is matched to up to *10* controls.
- Matching is restricted to individuals within the same time-indexed risk set.
- Exact matching constraints are applied on key variables (e.g. age band, sex, local authority district of residence, and length-of-stay/utilisation category), where specified.
- Propensity score distance is used to identify the closest matches among eligible controls.

This step generates matched sets linking each treated participant to one or more comparable controls.

##### (vii) Step 7. Derivation of cost components

Healthcare utilisation and cost outcomes are derived for the matched cohort across multiple domains:

- secondary care (inpatient, emergency department, outpatient),
- primary care (clinical and administrative contacts),
- prescribing and medication costs, and
- social care (where data completeness permits).

For each domain:

- utilisation events are extracted from the relevant data sources,
- costing rules or tariffs are applied, and
- costs are aggregated to the individual level over the follow-up period.

Each domain produces a separate cost table aligned to the matched cohort.

##### (viii) Step 8. Construction of final analysis dataset

All components are combined into a single analysis dataset:

- matched cohort structure and identifiers,
- baseline covariates,
- primary and secondary outcomes (including DAOH), and
- cost variables across all domains.

This final dataset forms the basis for:

- comparative effectiveness analyses, and
- health-economic evaluation.

